# High preoperative CEA and Systemic Inflammation Response Index(C-SIRI) Predict Unfavorable Survival of Resectable Colorectal Cancer

**DOI:** 10.1101/2023.04.11.23288397

**Authors:** Hao Cai, Yu Chen, Qiao Zhang, Yang Liu, HouJun Jia

## Abstract

**Background:** CEA and systemic inflammation were reported to correlate with proliferation, invasion and metastasis of colorectal cancer. This study investigated the prognostic significance of the preoperative CEA and systemic inflammation response index (C-SIRI) in patients with resectable colorectal cancer.

**Methods:** 217 CRC patients were recruited from Chongqing Medical University, the first affiliated hospital, between January 2015 and December 2017. Baseline characteristics, preoperative CEA level and peripheral monocyte, neutrophil and lymphocyte counts were retrospectively reviewed. The optimal cutoff value for SIRI was defined as 1.1, and for CEA, the best cutoff values were 4.1 ng/l and 13.0 ng/l. Patients with low levels of CEA (<4.1 ng/l) and SIRI (<1.1) were assigned a value of 0, those with high levels of CEA (≥13.0 ng/l) and SIRI (≥1.1) were assigned a value of 3, Those with CEA in the (4.1-13.0 ng/l) and SIRI (≥1.1), CEA (≥13.0 ng/l) and SIRI (<1.1) were assigned a value of 2. Those with CEA (<4.1 ng/l) and SIRI (≥1.1), CEA in the (4.1-13.0 ng/l) and SIRI (<1.1) were assigned a value of 1. The prognostic value was assessed based on univariate and multivariate survival analysis.

**Results:** Preoperative C-SIRI was significantly correlated with gender, site, stage, CEA, OPNI, NLR, PLR, and MLR. However, no difference was observed between C-SIRI and age, BMI, family history of cancer, adjuvant therapy, and AGR groups. Among these indicators, the correlation between PLR and NLR is the strongest. In addition, high preoperative C-SIRI was significantly correlated with poorer overall survival (OS) (HR: 2.782, 95%CI: 1.630-4.746, P<0.001) based on univariate survival analysis. Moreover, it remained an independent predictor for OS (HR: 2.563, 95%CI: 1.419-4.628, p=0.002) in multivariate Cox regression analysis.

**Conclusion:** Our study showed that preoperative C-SIRI could serve as a significant prognostic biomarker in patients with resectable colorectal cancer.

## Background

Globally, colorectal cancer (CRC) has become the third most common cancer and the second leading cause of tumor-related death. CRC accounts for approximately 10% of all new cancer diagnoses and cancer-related deaths each year [1,2]. Although recent advances in pathophysiological research have provided more treatment options and personalized treatment regimens and significantly improved overall survival in patients with advanced disease, CRC is still responsible for nearly 900,000 deaths per year [2]. Therefore, it is necessary to develop comprehensive indicators to evaluate the prognosis of patients.

Malignancy is the product of multiple steps in which the acquisition of specific capabilities, such as evading growth suppressors and resisting cell death, ultimately combine to contribute to cancer’s growth, invasion and metastasis. Among the contributing factors, tumor-associated inflammation is considered the seventh hallmark of cancer. Various factors, including smoking, chronic infections, environmental exposure such as silica and asbestos, and dietary habits, can lead to chronic inflammatory states in the host [3-4]. Inflammatory cells can release some chemicals, especially reactive oxygen species, which prompt the genetic evolution of surrounding cancer cells in a highly malignant direction [5]. From an etiological standpoint, these chronic inflammatory states lead to host genetic and epigenetic alterations, resulting in 25% of malignancy cases [6]. Moreover, local and systemic inflammation leads to the release of biochemicals, including survival factors that limit cell death, certain enzymes that facilitate angiogenesis and growth factors that sustain proliferative signaling, further promoting tumor progression [5,7,28]. Therefore, tumor-associated inflammation significantly impacts CRC patients’ survival at various levels of biology.

Specific preoperative biomarkers can be utilized to evaluate systemic inflammation in CRC patients. Previous studies have explored the prognostic significance of various indicators, such as neutrophil-to-lymphocyte ratio (NLR), lymphocyte-to-monocyte ratio (LMR), platelet-to-lymphocyte ratio (PLR), and C-reactive protein (CRP). In the preoperative phase, elevated levels of these biomarkers have been significantly associated with poorer survival [8-10]. In recent years, a novel indicator has been proposed to assess systemic inflammation. The systemic inflammation response index (SIRI) was established based on neutrophils, monocyte and lymphocyte. To date, the prognostic value of SIRI has been confirmed for various malignancies, such as pancreatic, cervical and gastric cancers, with limited evidence indicating a significant association with survival outcomes in CRC patients [11-14]. In addition, carcinoembryonic antigen (CEA) is a common tool used to evaluate the prognosis of CRC, and high preoperative CEA levels predict poor disease-free survival (DFS), overall survival (OS) and increased risk of recurrence and metastasis, as shown in earlier studies [15-16].

Therefore, we proposed a novel prognostic index based on CEA and SIRI (C-SIRI) to investigate whether it could accurately predict long-term survival in resectable CRC cases. Our aim was also to validate the prognostic value of SIRI in CRC patients and to provide research evidence for individual prediction and decision-making.

## Material and method

### Patients

From January 2015 to January 2017, a total of 300 CRC patients who underwent radical resection were consecutively enrolled at the First Affiliated Hospital of Chongqing Medical University (Chongqing, People’s Republic of China). The exclusion criteria were as follows: 1) people with a history of other primary malignant tumors or concurrent secondary malignancies, 2) neoadjuvant radiotherapy or radiochemotherapy ahead of surgery, 3) people with blood system diseases, infections, treatments which influence the biomarkers, 4) people directly or indirectly die of diseases other than CRC, 5) people with emergent surgeries precede inclusion, 6) people with other serious diseases which have a great influence on life expectancy, such as Intracerebral haemorrhage, myocardial infarction. Finally, 217 cases were included based on the criteria above. The study got approved by the independent Ethics Committee at The First Affiliated Hospital of Chongqing Medical University (K2023-104) and was conducted in line with the ethical standards of the World Medical Association Declaration of Helsinki.

### Data collection

The clinical information and blood indicators were obtained within 3days prior to surgery, such as age, gender, past medical history, smoking and drinking history, family history, BMI, neutrophil, lymphocyte, monocyte, albumin, carcinoembryonic antigen (CEA). Then some composite metrics were calculated according to the following formulas:albumin-to-globulin ratio (AGR), neutrphil×monocyte-to-lymphocyte ratio (SIRI, Systemic Inflammation Response Index), serum albumin+5×lymphocyte (OPNI/PNI, Onodera’s Prognostic Nutritional Index), neutrophil-to-lymphocyte ratio (NLR), monocyte-to-lymphocyte ratio (MLR).

### Follow-up and treatment

For patients at high risk of recurrence and metastasis, adjuvant chemotherapy was offered based on the patient’s wishes. Trained subject members followed up all patients via telephone. Overall survival(OS)was defined as the period from pathological confirmation of cancer to the patient’s death or the most recent follow-up.

### Definition of CEA and SIRI index

SIRI was calculated by the following equation:neutrophil×monocyte-to-lymphocyte ratio, and the optimal cutoff values for SIRI and CEA were obtained with x-tiles 3.6.1 software (Yale University, New Haven, CT, USA). Patients with low levels of CEA (<4.1 ng/l) and SIRI (<1.1) were assigned a value of 0, those with high levels of CEA (≥13.0 ng/l)and SIRI (≥1.1) were assigned a value of 3, Those with CEA in the (4.1-13.0 ng/l) and SIRI (≥1.1), CEA(≥13.0 ng/l) and SIRI (<1.1) were assigned a value of 2. Those with CEA (<4.1 ng/l) and SIRI (≥1.1), CEA in the (4.1-13.0 ng/l) and SIRI (<1.1) were assigned a value of 1.

### Statistical analysis

The optimal cutoff values of NLR, PLR, OPNI, AGR, MLR, and age were obtained by x-tiles software. Pearson’s χ2 test was utilized to reveal the correlation between variables. To find ind ependent factors in the prognosis of colorectal cancer, hazard ratios (HRs) and 95% confidence intervals (CIs) were evaluated based on the univariate and multivariate Cox regression model. P-value less than 0.05 was statistical significance. All statistical analyses were carried out using SPSS 22.0 (SPSS IBM, Chicago, IL, USA).

## RESULTS

### Patients’ characteristics

The study enrolled two hundred and seventeen patients (Table 1). The cohort included 124 (57. 1%) male and 93 (42.9%) female patients. Based on the 8th edition of the AJCC staging criteria, most patients (76.5%) were diagnosed with stage II and III diseases, which accounted for an enormous proportion (90.7%) of the population with outcome events. During the follow-up period, a total of 54 patients had outcome events.

**Table 1.**
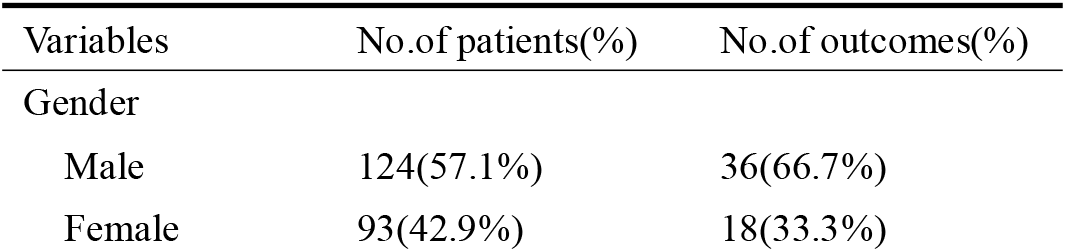
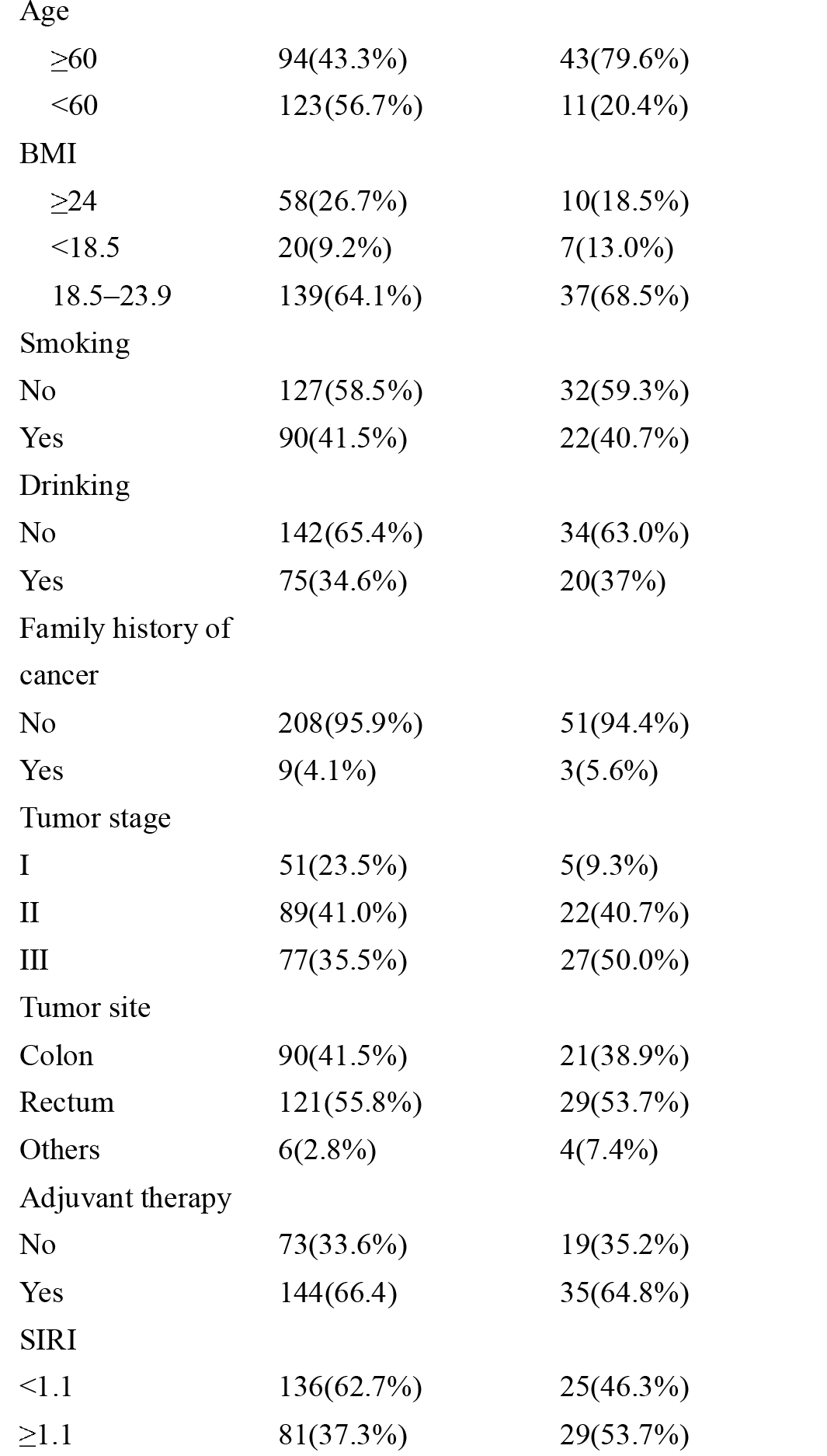
Baseline clinicopathological characteristics of patients with CRC

| Variables | No.of patients(%) | No.of outcomes(%) |
| --- | --- | --- |
| Gender |  |  |
| Male | 124(57.1%) | 36(66.7%) |
| Female | 93(42.9%) | 18(33.3%) |
| Age |  |  |
| ≥60 | 94(43.3%) | 43(79.6%) |
| <60 | 123(56.7%) | 11(20.4%) |
| BMI |  |  |
| ≥24 | 58(26.7%) | 10(18.5%) |
| <18.5 | 20(9.2%) | 7(13.0%) |
| 18.5–23.9 | 139(64.1%) | 37(68.5%) |
| Smoking |  |  |
| No | 127(58.5%) | 32(59.3%) |
| Yes | 90(41.5%) | 22(40.7%) |
| Drinking |  |  |
| No | 142(65.4%) | 34(63.0%) |
| Yes | 75(34.6%) | 20(37%) |
| Family history of cancer |  |  |
| No | 208(95.9%) | 51(94.4%) |
| Yes | 9(4.1%) | 3(5.6%) |
| Tumor stage |  |  |
| I | 51(23.5%) | 5(9.3%) |
| II | 89(41.0%) | 22(40.7%) |
| III | 77(35.5%) | 27(50.0%) |
| Tumor site |  |  |
| Colon | 90(41.5%) | 21(38.9%) |
| Rectum | 121(55.8%) | 29(53.7%) |
| Others | 6(2.8%) | 4(7.4%) |
| Adjuvant therapy |  |  |
| No | 73(33.6%) | 19(35.2%) |
| Yes | 144(66.4%) | 35(64.8%) |
| SIRI |  |  |
| <1.1 | 136(62.7%) | 25(46.3%) |
| ≥1.1 | 81(37.3%) | 29(53.7%) |

### Correlation between preoperative SIRI and other variables

Based on Pearson’s χ2 test, preoperative C-SIRI is correlated with gender, site, stage, CEA, OPNI, NLR, PLR, MLR (p=0.019, p=0.002, p=0.009, p<0.001, p=0.06, p<0.001, p=0.001 and p<0.001, respectively). However, there was no significant difference in age, BMI, family history of cancer, adjuvant therapy groups. The heat map provided the correlation between eight indicators, reflecting the value and the shade of color (**Fig.1**). Some strong correlations were observed. Among eight indicators, the correlation between PLR and NLR is the strongest.

**Fig. 1.**
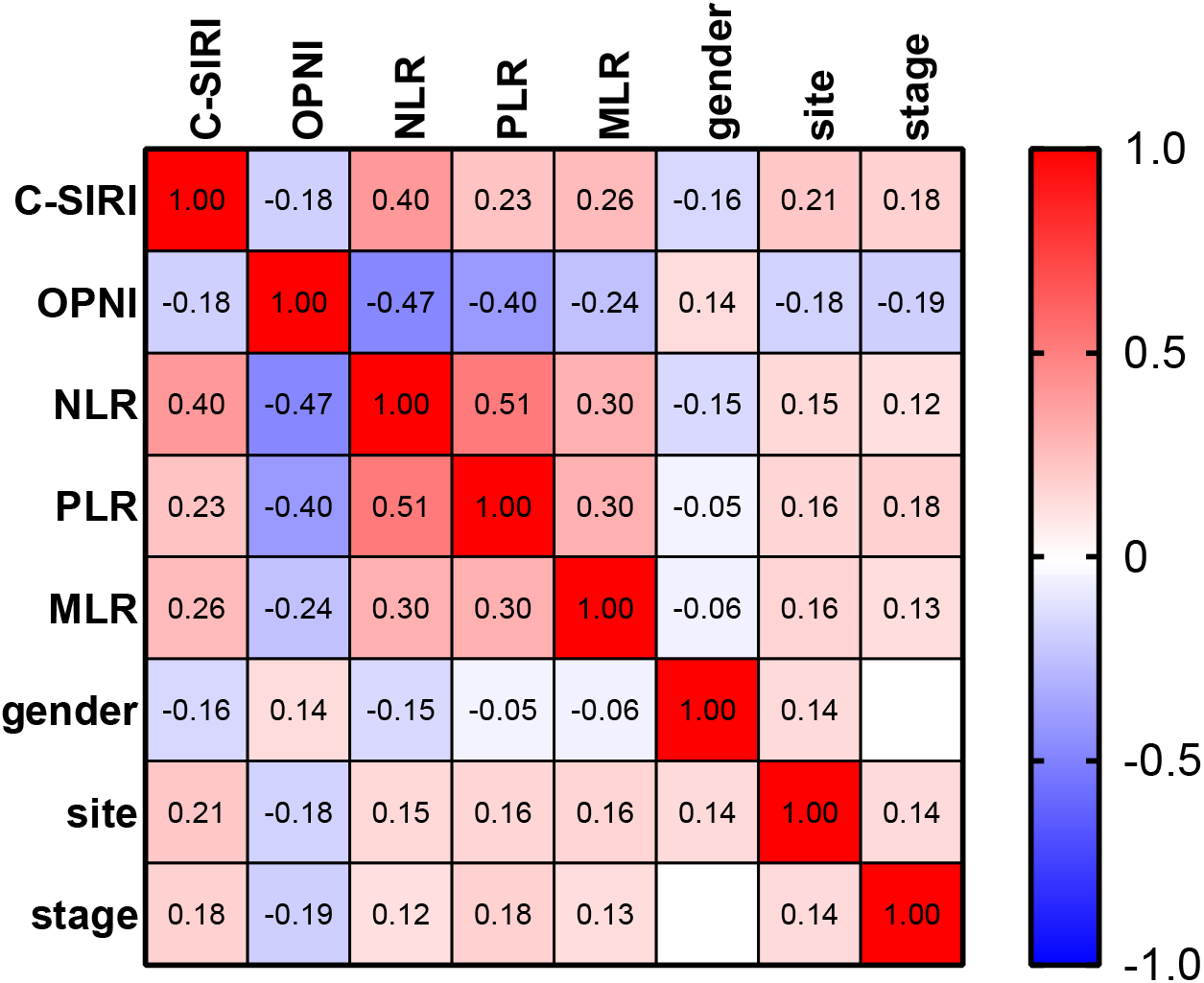
Correlation heat map of inflammation indicators.

### Prognostic Impact of Preoperative CEA or SIRI

The patients with low CEA value (<4.1 ng/l) had a better OS than those with high CEA level (4.1-13.0 ng/l or >13.0 ng/l) (p=0.016); **Fig.2B**). Similarly, low SIRI level was associated with prolonged overall survival in resectable colorectal cancer patients (p=0.02; **Fig.2A**).

**Fig. 2.**
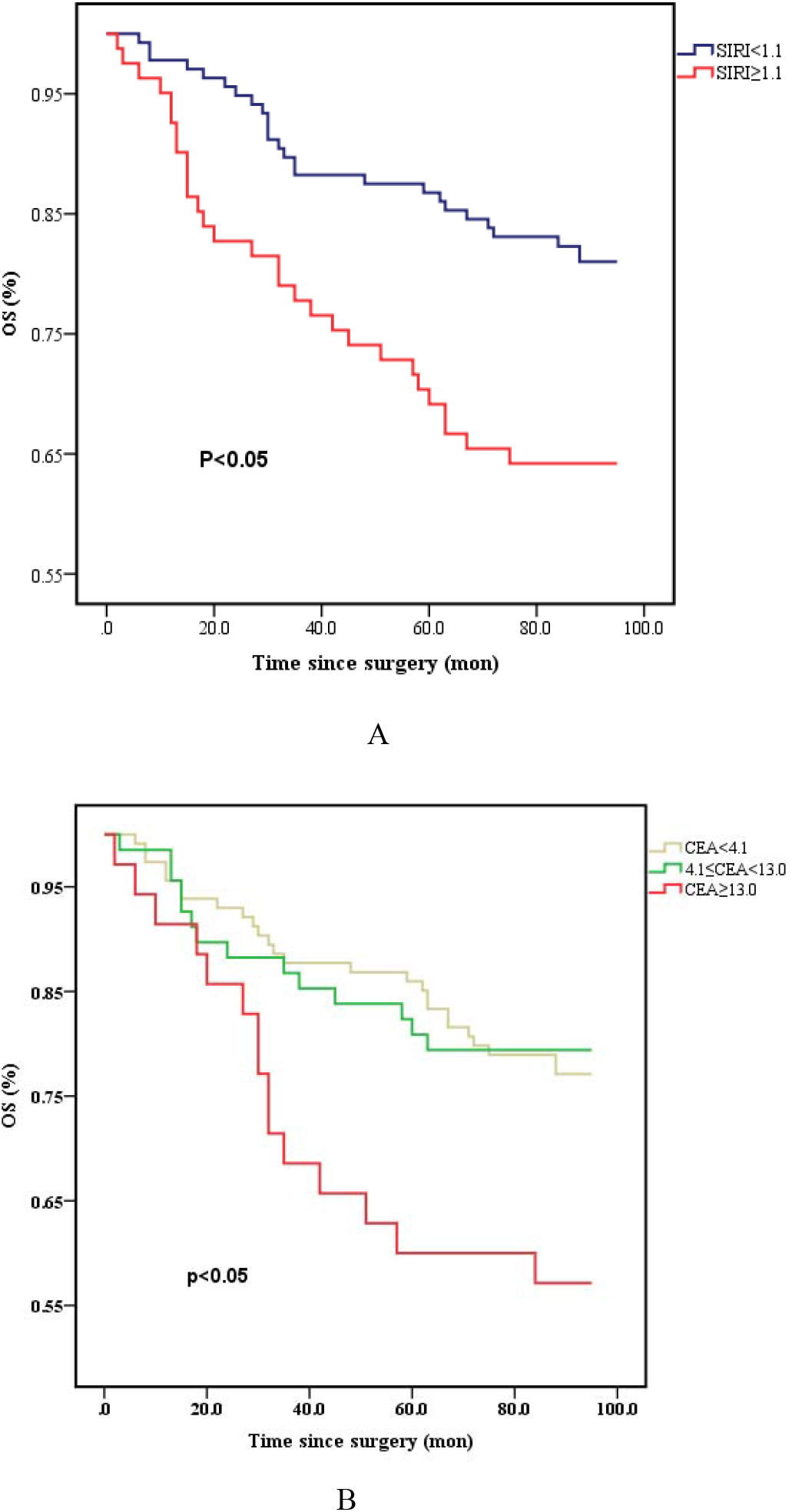
Kaplan-Meier survival curves of OS stratified by preoperative CEA levels (A) and SIRI (B) in 217 resectable colorectal cancer patients (with log-rank test).

### Prognostic significance of preoperative C-SIRI in resectable colorectal cancer

The prognostic value of C-SIRI compared with their higher counterparts, patients with lower C-SIRI tended to have a significantly better OS (p=0.002, **Fig.3**). The univariate and multivariate Cox models both showed that high C-SIRI was associated with an increased risk of OS. The HRs were 2.782 (95% CI: 1.630-4.746, P<0.001) and 2.563 (95% CI: 1.419-4.628, P=0.002), respectively. Additionally, preoperative C-SIRI (2/3 vs 0/1, HR: 2.563, 95%CI: 1.419-4.628, P= 0.002) remained an independent prognostic indicator for OS, based on multivariate analysis. At the same time, independent prognostic value for OS was found in age (<60 vs ≥60 HR: 0.30 0, 95%CI: 0.150-0.600) and tumor stage (III vs I stage, HR:5.392, 95%CI: 2.013-14.444). Furthermore, other parameters, including site, CEA, and SIRI, could also significantly predict OS (**Table 2**).

**Fig. 3.**
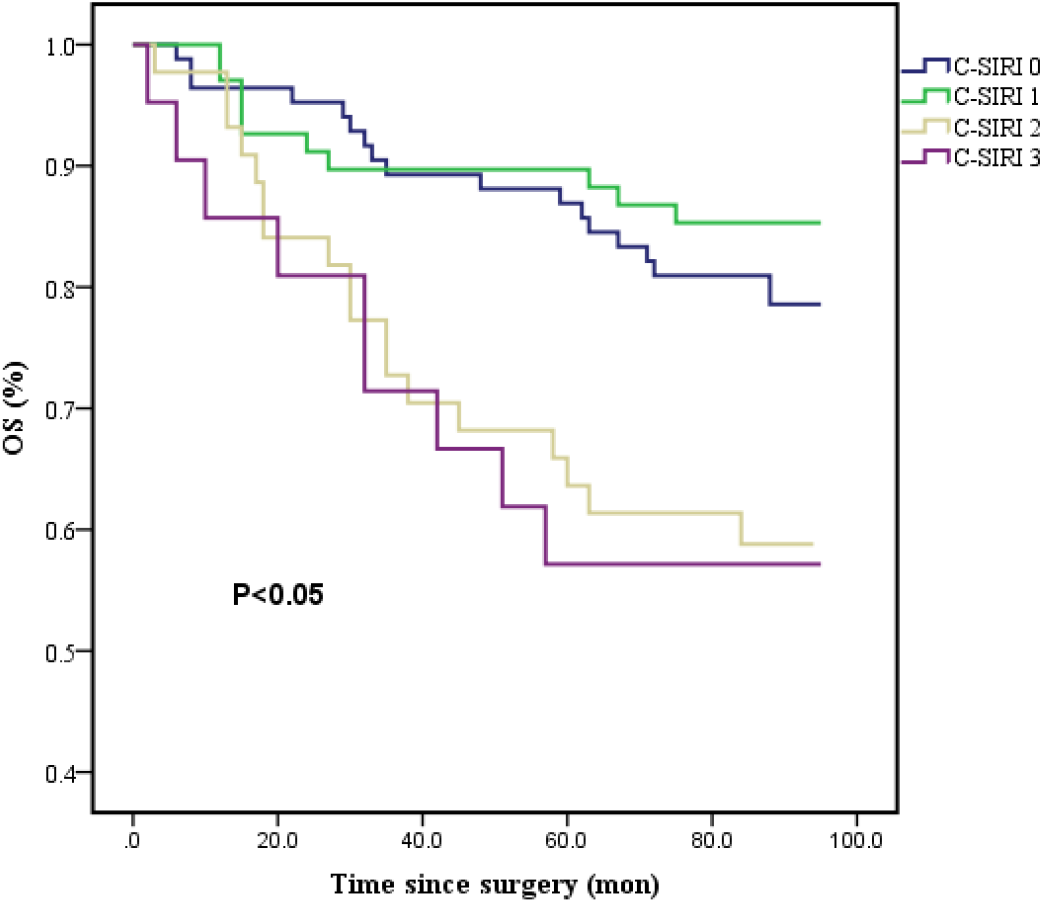
Kaplan-Meier survival curves of OS stratified by preoperative C-SIRI in 217 resectable colorectal cancer patients (with log-rank test).

**Table 2.**
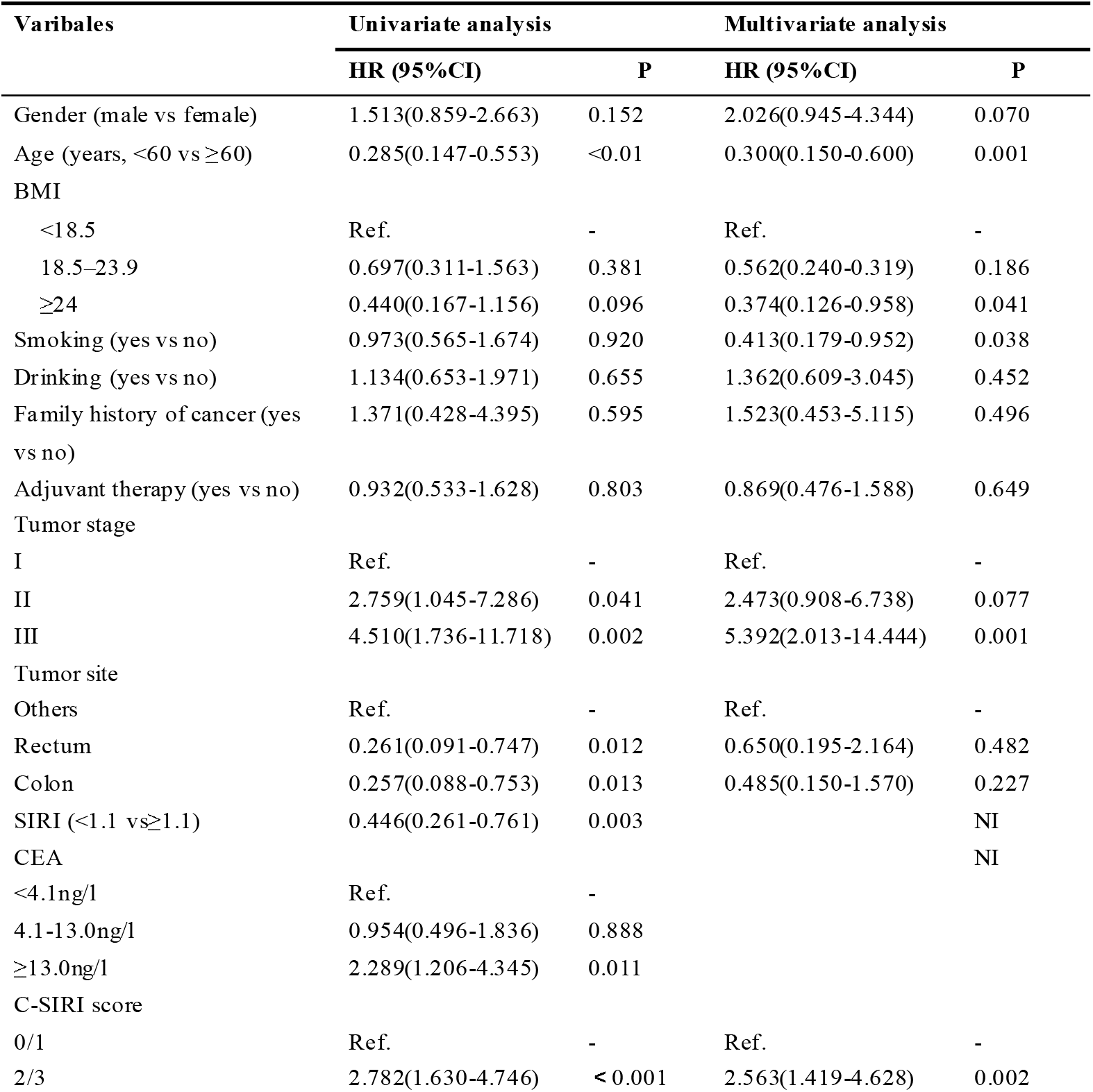
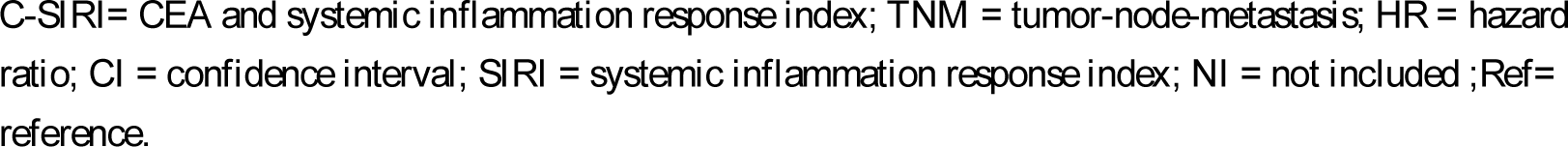
Univariate and multivariate analysis for overall survival.

| Varibales | Univariate analysis |  | Multivariate analysis |  |
| --- | --- | --- | --- | --- |
|  | HR (95%CI) | P | HR (95%CI) | P |
| Gender (male vs female) | 1.513(0.859-2.663) | 0.152 | 2.026(0.945-4.344) | 0.070 |
| Age (years, <60 vs ≥60) | 0.285(0.147-0.553) | <0.01 | 0.300(0.150-0.600) | 0.001 |
| BMI |  |  |  |  |
| <18.5 | Ref. | - | Ref. | - |
| 18.5–23.9 | 0.697(0.311-1.563) | 0.381 | 0.562(0.240-0.319) | 0.186 |
| ≥24 | 0.440(0.167-1.156) | 0.096 | 0.374(0.126-0.958) | 0.041 |
| Smoking (yes vs no) | 0.973(0.565-1.674) | 0.920 | 0.413(0.179-0.952) | 0.038 |
| Drinking (yes vs no) | 1.134(0.653-1.971) | 0.655 | 1.362(0.609-3.045) | 0.452 |
| Family history of cancer (yes vs no) | 1.371(0.428-4.395) | 0.595 | 1.523(0.453-5.115) | 0.496 |
| Adjuvant therapy (yes vs no) | 0.932(0.533-1.628) | 0.803 | 0.869(0.476-1.588) | 0.649 |
| Tumor stage |  |  |  |  |
| I | Ref. | - | Ref. | - |
| II | 2.759(1.045-7.286) | 0.041 | 2.473(0.908-6.738) | 0.077 |
| III | 4.510(1.736-11.718) | 0.002 | 5.392(2.013-14.444) | 0.001 |
| Tumor site |  |  |  |  |
| Others | Ref. | - | Ref. | - |
| Rectum | 0.261(0.091-0.747) | 0.012 | 0.650(0.195-2.164) | 0.482 |
| Colon | 0.257(0.088-0.753) | 0.013 | 0.485(0.150-1.570) | 0.227 |
| SIRI (<1.1 vs ≥1.1) | 0.446(0.261-0.761) | 0.003 |  | NI |
| CEA |  |  |  | NI |
| <4.1ng/l | Ref. | - |  |  |
| 4.1-13.0ng/l | 0.954(0.496-1.836) | 0.888 |  |  |
| ≥13.0ng/l | 2.289(1.206-4.345) | 0.011 |  |  |
| C-SIRI score |  |  |  |  |
| 0/1 | Ref. | - | Ref. | - |
| 2/3 | 2.782(1.630-4.746) | <0.001 | 2.563(1.419-4.628) | 0.002 |

C-SIRI= CEA and systemic inflammation response index; TNM = tumor-node-metastasis; HR = hazard ratio; CI = confidence interval; SIRI = systemic inflammation response index; NI = not included ;Ref= reference.

## Discussion

To the best of our knowledge, the present study is one of few studies to establish a combined indicator that unites CEA and SIRI in resectable colorectal cancer. Our study showed that preoperative C-SIRI was significantly associated with clinical outcomes and possessed independent prognostic value. Patients with high preoperative C-SIRI had a poorer prognosis. Additionally, preoperative SIRI has predictive value for the survival of CRC patients.

Recent studies have highlighted the important role of systemic inflammation in tumor growth, proliferation, metastasis and survival. It is a promising new direction for tumor treatment and monitoring. Several indicators have been reported to assess the level of systemic inflammation, such as NLR, PLR, MLR, LMR, and SIRI, and have been shown to have prognostic values in multiple tumors [17, 18]. Notably, SIRI was first proposed by Qi et al. as an independent prognostic marker in patients with pancreatic cancer [12]. Subsequent studies have affirmed the association of SIRI with survival in patients with liver, gastric, nasopharyngeal, and breast cancers [19-21]. Moreover, Cao et al. found that high preoperative SIRI was significantly associated with poorer OS and DFS of CRC patients, and predictive ability for the survival of CRC patients was superior to other inflammatory biomarkers, such as PLR, NLR, and SII [14]. However, to date, However, to date, multi-center data still need to confirm its relationship with CRC patients’ survival adequately.

In addition, CEA has first identified in fetal gut tissue and circulatory system of CRC patients over 50 years ago [22]. Subsequently, CEA was detected in the tumors from the gastrointestinal tract [23]. Despite the limited value of CEA for CRC screening, as elevated CEA levels may be due to some non-malignant diseases such as chronic inflammatory bowel disease, pancreatitis and liver disease [24], substantial evidence has confirmed its predictive ability in the recurrence, metastasis, and survival of CRC patients. Becerra AZ and his colleagues found that elevated preoperative CEA levels were associated with a 62% increased risk of death compared to normal CEA levels [25], and the 5-year overall survival was 74.5% vs 63.4% [26]. Besides, Kim et al. suggested that elevated CEA level was expected to decrease exponentially after curative surgery, and survival was significantly better than that of patients with a sustained high level of CEA [27].

Therefore, we proposed that C-SIRI might predict accurately in resectable CRC patients. Our study results suggest that C-SIRI is significantly correlated with gender, site, stage, CEA., OPNI, NLR, PLR and MLR. Among these factors, the strongest correlation was observed between PLR and NLR. However, no correlation was found between C-SIRI and BMI, adjuvant therapy, or AGR, indicating a need for further investigation. In addition, the study found that SIRI and CEA were both significant prognostic factors based on univariate analysis. Multivariate analysis revealed that C-SIRI remained an independent prognostic predictor for OS (HR, 2.563, 95% CI,1.419-4.628, p=0.002). Moreover, compared to the CRC patients with a low level of SIRI, those with SIRI ≥1.1 had a poorer prognostic outcome after curative resection, which is consistent with the prognostic value of SIRI in other malignancies. Thus, C-SIRI can serve as a reliable prognostic biomarker to support preoperative systemic inflammation response assessment and predict survival in CRC patients.

Although the present study has some limitations which should be considered, it provides valuable insights into the prognostic value of the novel indicator C-SIRI. Firstly, due to the limitation of specimen acquisition, the relationship between the local inflammation response of tumor lesions and systemic inflammatory response and its prognostic value were not investigated. Secondly, as a single-center retrospective study, potential selection bias may exist. Thirdly, the small sample size calls for more research to support the findings. Therefore, multi-center studies with large samples and external validation are necessary for the future.

## Conclusion

In conclusion, the study showed that C-SIRI was significantly associated with OS in CRC patients and confirmed its prognostic value based on univariate and multivariate analysis. This supports more accurate risk assessment and personalized treatment for resectable CRC patients.

## Data Availability

All data produced in the present study are available upon reasonable request to the authors.

## Authors’ contribution

HC was responsible for obtaining and analyzing data, drafting manuscripts, and making critical revisions. YC was responsible for patient follow-up and related clinicopathological data collection. QZ and YL were responsible for technical support for data analysis, article grammar proofreading. HJJ was responsible for the conception, design, and review of selected topics.

## Funding

The author(s) reported that there is no funding associated with the work featured in this article.

### Acknowledgement

None.

## Availability of data and materials

The datasets used in this study are available on request from the corresponding author.

## Declaration

### Ethics approval and consent to participate

This study was approved by the ethics committee of Chongqing Medical University (K2023-10 4) and followed the ethical standards of the Helsinki Declaration.

### Consent to participate

Not applicable.

